# First-in-Human Trial of a Recombinant Stabilized Prefusion SARS-CoV-2 Spike Protein Vaccine with Adjuvant of Aluminum Hydroxide and CpG 1018

**DOI:** 10.1101/2021.03.31.21254668

**Authors:** Szu-Min Hsieh, Wang-Da Liu, Yu-Shan Huang, Yi-Jiun Lin, Erh-Fang Hsieh, Wei-Cheng Lian, Charles Chen, I-Chen Tai, Shan-Chwen Chang

**Affiliations:** Section of Infectious Diseases, Division of Infectious Diseases, Department of Internal Medicine, National Taiwan University Hospital and College of Medicine, National Taiwan University, Taiwan; Medigen Vaccine Biologics Corp., Taiwan; College of Science and Technology, Temple University, Philadelphia, PA 19122, U.S.A.

**Author notes:** Corresponding authors I-Chen Tai, Corresponding Author, Postal address: Medigen Vaccine Biologics Corp., 7F., No. 16, Ln. 120, Sec. 1, Neihu Rd., Taipei 114, Taiwan, #604, Shan-Chwen Chang, Corresponding Author, Postal address: National Taiwan University Hospital, No.7, Zhongshan S. Rd., Zhongzheng Dist., Taipei City 100, Taiwan, #887999.

**Keywords:** COVID-19 vaccine, CpG 1018, S-2P protein

## Abstract

**Design:** This is a phase 1, dose-escalation open-label trial to evaluate the safety and immunogenicity of MVC-COV1901, a recombinant stabilized prefusion SARS-CoV-2 spike (S-2P) protein vaccine with adjuvant of aluminum hydroxide and CpG 1018.

**Methods:** We enrolled 45 healthy adults from 20 to 49 years of age to be administered with two vaccinations of MVC-COV1901 in a low dose (LD), middle dose (MD), and high dose (HD) of spike protein at 28 days apart. There were 15 participants in each dose group, and all of them were followed up for 28 days after the second vaccination at the time of interim analysis. Adverse events (AEs) and laboratory data were recorded for safety evaluation. Blood samples were collected for wild-type SARS-CoV-2 and pseudovirus neutralization assays as well as SARS-CoV-2 spike-specific immunoglobulin G (IgG) at various times. Overall, the study duration will be 7 months.

**Results:** Solicited events were mostly mild and similar in the participants of all three dose groups. No subject experienced fever. There were no serious nor adverse events of special interest at the time point of this interim report. After the second vaccination, the SARS-CoV-2 spike specific IgG titers increased with peak geometric mean titers at 7178.245 (LD), 7746.086 (MD), and 11220.58 (HD), respectively. Serum neutralizing activity was detected by two methods in all participants of MD and HD groups, with geometric mean values generally comparable to those of a panel of control convalescent serum specimens. All of the participants in the MD and HD groups were seroconverted after the second vaccination.

**Conclusions:** The MVC-COV1901 vaccine is safe and elicits remarkable immune responses especially in the MD and HD groups.

**Trial Registration:** ClinicalTrials.gov NCT 04487210

## Introduction

Human infections due to SARS-CoV-2 began to spread globally following the outbreak in Wuhan, China. WHO panel declared the COVID-19 outbreak as a public health emergency of international concerns, and this outbreak was also subsequently characterized as a pandemic declared by the WHO on March 11^th^, 2020.^1^

Approximately 15% of COVID-19 cases are severe that requires oxygen support, and 5% are critical disease with complications such as respiratory failure, acute respiratory distress syndrome (ARDS), sepsis, septic shock, etc.^2^ A meta-analysis assessed the risk of underlying diseases in severe patients compared to those in non-severe conditions and showed the pooled odds ratios were 2.36 for hypertension, 2.46 for respiratory system disease, and 3.42 for cardiovascular disease and 2.07 for diabetes.^3^

There is currently no cure medication for the potentially lethal COVID-19. Development of a range of vaccines will provide flexibility with prevention strategies. For vaccines intended to generate protective immune response, using an antigen with proper conformation is critical. The neutralizing antibodies induced by spike (S) protein block viruses from binding to their target receptor ACE2 and hence inhibit viral infection. S protein has two major conformational stats, prefusion and postfusion.^4^ S-2P protein is a recombinant version of the S protein developed by the laboratory of Dr. Barney S. Graham (Vaccine Research Center, National Institute of Allergy and Infectious Diseases[NIAID], U.S.A.), and is a stabilized prefusion S ectodomain, encoding residues 1-1208 of SARS-CoV-2 spike protein with two proline substitutions at residues 986 and 987, a “GSAS” substitution at residues 682–685 to abolish the furin cleavage site, and a T4 fibritin trimerization motif at the C-terminus.^5^ The cryo–electron microscopy structure showed the protein produced by this construct is in the prefusion conformation and can bind to ACE2.^5(p1261)^ Similar strategy had been used to retain MERS-COV S protein in the prefusion conformation and demonstrated that stabilized MERS-COV S protein was able to elicit high neutralizing antibody.^6^ Medigen’s MVC-COV1901 vaccine is formulated as S-2P adjuvanted with Dynavax’s CpG 1018 and aluminum hydroxide. CpG 1018 is an oligodeoxynucleotide which acts as a toll-like receptor 9 agonist and has been shown in our preclinical studies to enhance immunogenicity and induce a Th1-skewed immune response^7^.

## Methods

### Trial Design

This study was a phase 1, prospective, open-label, dose-escalation study to evaluate the safety and immunogenicity of a SARS-CoV-2 vaccine in adults aged 20 to 49 years. The study was commenced at the National Taiwan University Hospital in northern Taiwan from September 2020. The trial protocol and informed consent form were approved by the Taiwan Food and Drug Administration and the ethics committee at the site. The trial was conducted in accordance with the principles of the Declaration of Helsinki and Good Clinical Practice. An independent data and safety monitoring board (DSMB) was established to monitor safety data. (ClinicalTrials.gov NCT 04487210)

### The Investigational Vaccine

We applied technology previously used for MERS-CoV to produce a prefusion-stabilized SARS-CoV-2 spike protein, S-2P, designed by Dr. Barney S. Graham (Vaccine Research Center, NIAID, U.S.A.). The MVC-COV1901 contained S-2P protein adjuvanted with CpG 1018 and aluminum hydroxide as previous reported.^7(p7)^. The vaccine administered at a volume of 0.5 mL as a single dose via intramuscular injection. The production of the vaccine was conducted in Medigen Vaccine Biologics Corp. facility which is compliant with the current good manufacturing practices (cGMP).

### Participants

Eligible participants were healthy adults from 20 to 49 years of age. Eligibility was determined based on medical history, physical examination, laboratory tests, and investigators’ clinical judgment. Exclusion criteria included a history of known potential exposure to SARS CoV-1 or 2 viruses, having received any other COVID-19 vaccine, impaired immune function, history of autoimmune disease, abnormal autoantibody tests, febrile or acute illness within 2 days of first dose, and acute respiratory illness within 14 days of first dose.

### Interventions

This study was a dose escalation study with three separate sub-phases for participants from 20 to 49 years of age. Each sub-phase consisted of 15 participants. The three different dose levels employed in this clinical trial are LD, MD, and HD of S-2P protein adjuvanted with CpG 1018 and aluminum hydroxide for phase 1a, 1b, and 1c, respectively. The vaccination schedule consisted of two doses, administered by intramuscular (IM) injection of 0.5 mL in the deltoid region of non-dominant arm 28 days apart, on Day 1 and Day 29. The protocol permitted an interim analysis to make decisions regarding vaccine strategy.

#### Phase 1a

Four sentinel participants would be recruited to receive LD of S-2P protein with adjuvant to evaluate preliminary safety data of the vaccine. If no ≥ Grade 3 AE or SAE occurred within 7 days after the first vaccination in the 4 sentinel participants, dosing of the remaining participants in Phase 1a and Phase 1b would proceed.

#### Phase 1b

Another 4 sentinel participants would be enrolled to receive MD of S-2P protein with adjuvant in Phase 1b. If no ≥ Grade 3 AE or SAE occurred within 7 days after the first vaccination in the 4 sentinel participants, dosing of the remaining participants in Phase 1b and Phase 1c would proceed.

#### Phase 1c

Another 4 sentinel participants would be enrolled to receive HD of S-2P protein with adjuvant in phase 1c. If no ≥ Grade 3 AE or SAE occurred within 7 days after the first vaccination in the 4 sentinel participants, dosing of the remaining participants in Phase 1c would proceed.

An interim analysis of immunogenicity data and safety data from baseline to 28 days after the second vaccination for all participants were carried out when all of the participants have completed the visit (at 28 days after the second vaccination).

### Safety Assessment

The primary endpoint was to evaluate the safety of MVC-COV1901 in three different strengths (LD, MD, and HD of S-2P protein adjuvanted with CpG 1018 and aluminum hydroxide) from Day 1 to 28 days after the second vaccination. Vital signs and electrocardiogram (ECG) were performed before and after vaccination. Participants were observed for at least 30 min after each vaccination to identify any immediate adverse events (AEs), and were asked to record solicited local and systemic AEs in the participant’s diary card for up to 7 days after each vaccination. Unsolicited AEs were recorded for 28 days following each vaccination; all other AEs, serious adverse events (SAEs) and adverse events of special interests (AESIs) were recorded throughout the study period.

Serum samples were collected for safety evaluations. Hematology, biochemistry and immunology tests were measured as well.

### Immunogenicity Analysis

The immunogenicity endpoints were to evaluate the immunogenicity in terms of neutralizing antibody titers and binding antibody titers at 14 days (Day 15) and 28 days (Day 29) after first and at 14 days (Day 43) and 28 days (Day 57) after second vaccination, as well as 90 days and 180 days after the second vaccination.

#### SARS-CoV-2 spike-specific immunoglobulin G (IgG)

Total serum anti-Spike IgG titers were detected with direct enzyme-linked immunosorbent assay (ELISA) using customized 96-well plates coated with S-2P antigen.

#### SARS-CoV-2 pseudovirus neutralization assay

Pseudovirus production and titration followed the previous report.^7(p7)^ Serial dilutions of the samples to be tested were performed (initial dilution of 1:20; diluted two-fold to a final dilution of 1:2560). The diluted serum was mixed with equal volume of pseudovirus (1000 TU) and incubated before adding to the plates with HEK293-hAce2 cells (1 × 10^4^ cells/well). The amount of pseudovirus entering the cells was calculated by lysing and measuring the relative luciferase units (RLU). Fifty percent inhibition dilution (concentration) titers (ID_50_) were calculated considering uninfected cells as 100% neutralization and cells transduced with virus as 0% neutralization and reciprocal ID_50_ geometric mean titers (GMT) were both determined.

#### Wild□type SARS□CoV□2 neutralization assay

SARS-CoV-2 virus (hCoV-19/Taiwan/CGMH-CGU-01/2020, GenBank accession MT192759) was obtained and titrated to obtain TCID_50_, and Vero E6 cells (2.5 × 10^4^ cells/well) were seeded in 96-well plates and incubated. The sera underwent a total of 11 two-fold dilutions with the final dilution being 1:8192, and the diluted sera were mixed with equal volume of viral solution containing 100 TCID_50_. The serum-virus mixture was incubated and then added to the cell plates containing the Vero E6 cells, followed by further incubation. The neutralizing titer was defined as the reciprocal of the highest dilution capable of inhibiting 50% of the CPE (NT_50_), which was calculated in accordance with the formula of the Reed-Muench method.

### Statistical Analysis

Safety analysis was performed on the total vaccinated group (TVG) population who received at least 1 dose of vaccine. The endpoints related to immunogenicity consisted of the following: antigen specific immunoglobulins, neutralizing antibody titers of wild type virus and pseudovirus, in terms of geometric mean titer (GMT) and seroconversion rate (SCR). SCR is defined as the percentage of participants with ≥ 4-fold increase in titers from the baseline or from half of the lower limit of detection (LoD) if undetectable at baseline.

## Results

### Study Population

In total, 77 participants were screened. Of these, 45 eligible participants had completed two doses of MVC-COV1901. Participants’ baseline demographic characteristics are summarized in Table 1.

**Table 1.** Demographic Characteristics of Eligible Participants.

| **Demographic characteristics** | **LD** | **MD** | **HD** | **Total** |
| --- | --- | --- | --- | --- |
| **No. of Participants** | 15 | 15 | 15 | 45 |
| **Age** |  |  |  |  |
| Mean (SD), years | 36.7 (8.97) | 33.3 (8.03) | 31.5 (5.78) | 33.8 (7.84) |
| **Gender** |  |  |  |  |
| Male, No. (%) | 7 (46.7%) | 9 (60.0%) | 12 (80.0%) | 28 (62.2%) |
| Female, No. (%) | 8 (53.3%) | 6 (40.0%) | 3 (20.0%) | 17 (37.8%) |
| **BMI (kg/m<sup>2</sup>)** |  |  |  |  |
| Mean (SD) | 23.18 (3.394) | 23.30 (3.084) | 23.17 (2.429) | 23.22 (2.928) |

### Vaccine Safety

Neither SAEs nor AESI occurred at this time point of interim analysis. No participants missed the second dose. All of the local and systemic AEs were mild, except for one malaise/fatigue in the HD group. None of the participants had fever after either dose 1 or dose 2. Solicited adverse events after the first and the second vaccination were similar. Evaluation of safety laboratory values, ECG interpretation, and unsolicited adverse events revealed no specific concern.

### Immunogenicity

The humoral immunogenicity results are summarized in Figure 1.

**Fig 1.**
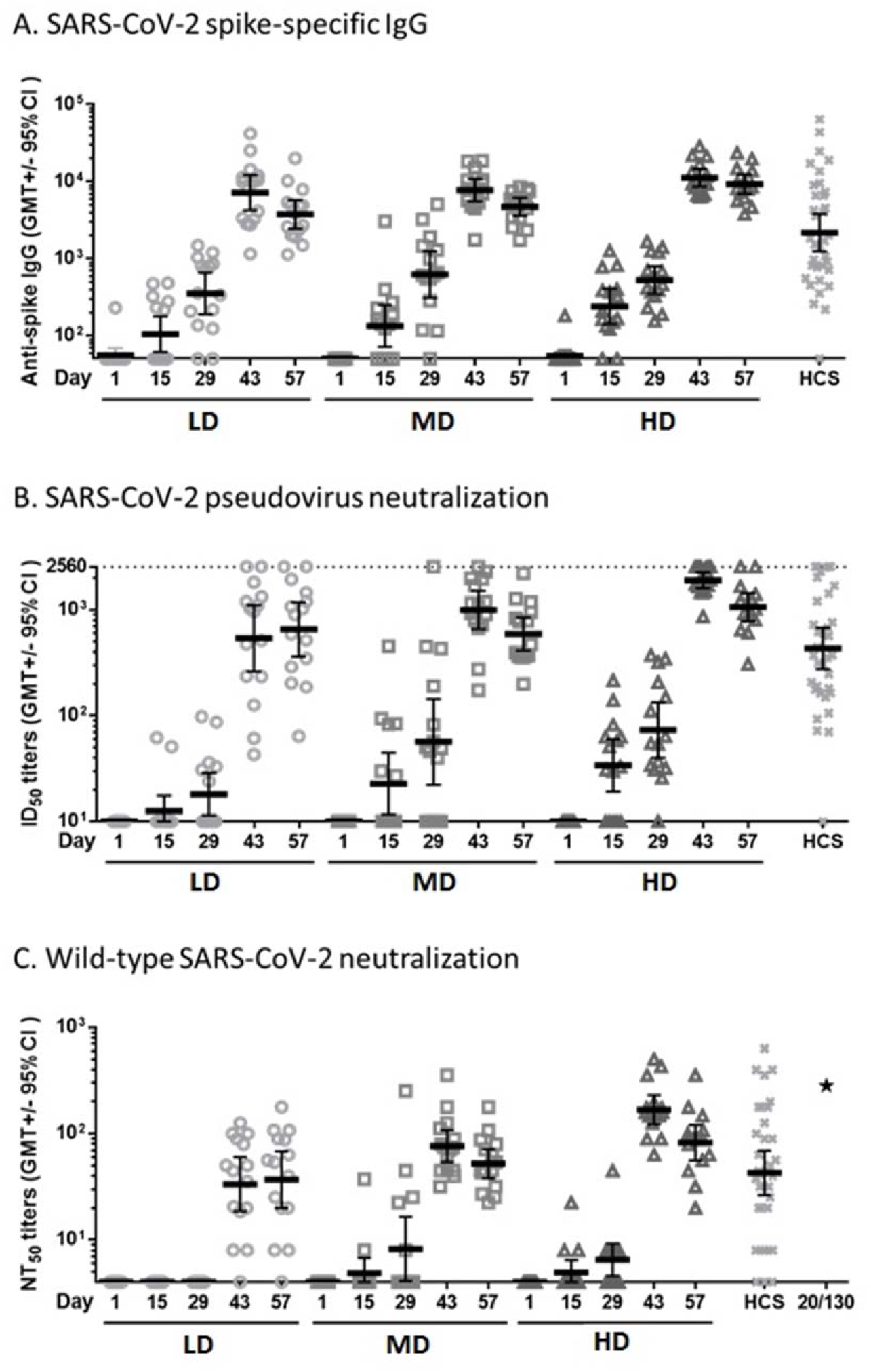
Summary of Humoral Immune Response. Sera of participants vaccinated with LD, MD, or HD of MVC-COV1901 were measured for anti-spike IgG by (A) ELISA, and neutralization titers were measured by (B) pseudovirus neutralization assay or (C) live virus neutralization assay. Human convalescent sera (HCS) from 35 recovered COVID-19 patients were analysed by the same assays for comparison and NIBSC 20/130 standard was used in the live virus neutralization assay as a standard (asterisk in panel C). Bars indicate geometric mean titers and error bars indicate 95% confidence intervals. A: The GMTs at Day 43 in terms of SARS-CoV-2 spike specific IgG were 7178.245 (LD), 7746.086 (MD), 11220.58 (HD), and 2179.598 (HCS); B: The GMTs at Day 43 in terms of SARS-CoV-2 pseudovirus ID_50_ were 538.496 (LD), 993.075 (MD), 1905.840 (HD), and 430.468 (HCS); C: The GMTs at Day 43 in terms of wild type SARS-CoV-2 NT_50_ were 33.317 (LD), 76.307 (MD), 167.402 (HD), and 42.674 (HCS). The value of NIBSC 20/130 standard was 281.84. (as shown in asterisk)

#### SARS-CoV-2 spike specific immunoglobulin G (IgG)

Binding IgG titers to S protein increased rapidly after the second vaccination, with seroconversion in all participants by Day 43 and 57. The GMTs peaked at Day 43 with value of 7178.245 (95% CI: 4240.336 ∼12151.68), 7746.086 (95% CI: 5530.230 ∼10849.79), 11220.58 (95% CI: 8592.293 ∼14652.84) in LD, MD, HD groups, respectively. The responses in LD, MD, and HD groups were also similar to the values of convalescent serum specimens.(2179.598, [95%CI: 1240.897∼3828.396]). (Fig 1A)

#### Neutralization responses for SARS-CoV-2 pseudovirus

Before vaccination, no subject had detectable pseudovirus neutralizing titers (ID_50_) at the lower limit of serum concentration tested (1:20 dilution) in the assay. At Day 43, the pseudovirus neutralizing titers (ID_50_) showed peaked GMTs of 538.496 (95% CI: 261.9403 ∼1107.036), 993.075 (95% CI: 654.9991 ∼1505.648), and 1905.840 (95% CI: 1601.667 ∼2267.779) in LD, MD, and HD groups, respectively. All of the participants in three doses were seroconverted after the second vaccination (Fig 1B).

#### Neutralization responses for wild type SARS-CoV-2 virus

Before vaccination, no subject had detectable wild-type virus neutralizing titers (NT_50_) at the lower limit of serum concentration tested (1:8 dilution) in the assay. However, after the second vaccination, neutralizing responses were identified in serum samples from all participants in MD and HD groups. At Day 43, the GMTs were 33.317 (95% CI: 18.5239 ∼59.9256), 76.307 (95% CI: 53.7497 ∼108.3309), and 167.402 (95% CI: 122.0492 ∼229.6090) in LD, MD, and HD groups, respectively. The MD and HD groups were similar in terms of GMT (52.205 [95% CI: 37.9448 ∼71.8246] and 81.909 [95% CI: 55.8131 ∼120.2070], respectively, at Day 57). These responses were also similar to the value of convalescent serum specimens. (42.674, [95%CI: 26.37666∼69.0403]) All participants in MD and HD groups were seroconverted at Day 43 and Day 57. The results of wild-type SARS-CoV-2 neutralizing antibody titer are summarized in Fig 1C.

## Discussion

This is the first clinical trial report to address the protein-based vaccine using the S-2P protein developed by NIAID, U.S.A. as the antigen, and adjuvanted with CpG 1018 and aluminum hydroxide.

This interim analysis demonstrated that the MVC-COV1901 vaccine was well tolerated and immunogenic in healthy adults aged 20 to 49 years. Across the three dose groups, local injection-site reactions were all mild. This safety profile is similar to that described in the previous report for subunit protein vaccines adjuvanted with CpG 1018 or aluminum hydroxide.^8, 9^ With regards to the occurrence rate and severity of solicited AEs, no differences were noticed among the LD, MD, and HD groups, or between the first and second vaccination.

The rates of detectable neutralizing response in all three dose groups at baseline were 0%, aligning with the fact that there was no circulating SARS-CoV-2 in Taiwan. The neutralizing antibody titers were measurable at Day 43 and Day 57 for all dose levels. All participants in MD and HD groups were seroconverted in terms of wild-type SARS-CoV-2 neutralizing response. Besides, the wild type neutralizing antibody response profile also demonstrated a good correlation with IgG and pseudovirus neutralizing antibody titers. Although the correlation of protection is not available at present, serum neutralizing activity has been shown to be correlate of protection for other viruses while developing vaccines, such as yellow fever vaccine, polio vaccine, and Japanese encephalitis vaccine,^10^ and is generally accepted as a useful biomarker of the in vivo humoral response. Besides, the preclinical study of MVC-COV1901^11^ had shown that hamsters were protected from SARS-CoV-2 virus challenge after two vaccinations of S-2P protein adjuvanted with CpG and aluminum hydroxide. The geometric mean titers in the MD and HD groups were comparable with those of a panel of control convalescent serum specimens with all participants in both groups seroconverted after two vaccinations. Therefore, a MD of S-2P combined with CpG 1018 and aluminum hydroxide was deemed adequate to elicit a profound humoral immune response.

This interim report has some limitations: small size of the trial, the short period of follow-up at this time point, and the participants’ young age and good health status. We were not able to assess the durability of the immune responses after Day 57 in this interim report. However, participants will be followed up for 6 months after the second vaccination with scheduled blood collections throughout that period to evaluate the humoral immunologic responses. Although the level of immunity needed to protect from COVID-19 remains unknown, NIBSC 20/130 standard serum was tested in terms of wild-type SARS-CoV-2 neutralizing antibody titer for the development and evaluation of serological assays for the detection of antibodies against SARS-CoV-2, as a positive control.

These safety and immunogenicity findings support further advancement of the MVC-COV1901 vaccine to subsequent clinical trials. Of the three doses evaluated, both the MD and HD elicited high neutralizing antibody responses with all participants seroconverted after second vaccination. Further phase 2 trial with 3700 participants (including the populations at greatest risk for serious Covid-19 such as those with chronic medical diseases and older adults) is on-going (ClinicalTrials.gov number, NCT04695652)

## Data Availability

The datasets generated during and analyzed during the current study are available from the corresponding author on reasonable and expressively written request.

## Author contributions

SM Hsieh, SC Chang and IC Tai had full access to all of the data in the study and take responsibility for the integrity of the data and the accuracy of the data analysis. SC Chang and IC Tai contributed equally and are joint corresponding authors.

Concept and design: SM Hsieh and IC Tai

Acquisition, or interpretation of data: SM Hsieh, SC Chang, WD Liu, and YS Huang Drafting and preparing the manuscript: SM Hsieh, SC Chang, WD Liu, YS Huang, YJ Lin, EF Hsieh, and IC Tai

Critical revision of the manuscript for important intellectual content: SM Hsieh, SC Chang Laboratory assays set up and analysis of data: YJ Lin, and EF Hsieh

Administrative, technical, or material support: Charles Chen and WC Lian

All authors reviewed and approved of the final version of the manuscript.

## Additional Contributions

All the participants for their dedication to this trial; Dr. Barney S. Graham at Vaccine Research Center, National Institute of Allergy and Infectious Diseases, U.S.A., team members at Dynavax Technologies, Emeryville, CA 94608, USA. and Tsun□Yung Kuo at Department of Biotechnology and Animal Science, National Ilan University, Ilan, Taiwan for providing technical guidance and helpful advice; The investigational staff at National Taiwan University Hospital, Taiwan and A2 Healthcare Taiwan Corp. for the conduction of the trial; Leo Lee, Tzay Huar Hong, Hui-Yi Wang, Chian-En Lien, Hao-Yuan Cheng and at Medigen Vaccine Biologics Corp. for collaboration on protocol development, significant contribution to the Investigational New Drug (IND) application, and participation in weekly meeting with the regulatory authority; The members of the safety monitoring committee; Dr. Yu-Chi Chou and his team members at the RNAi Core Facility, Academia Sinica for the pseudovirus neutralization assay; Dr. Shin-Ru Shih at Chang Gung University, Taoyuan, Taiwan and Dr. Chung□Guei Huang as well as her team members at Chang Gung Memorial Hospital, Taoyuan, Taiwan for the wild type SARS-CoV-2 neutralization assay; Team members at Protech Pharmaservices Corporation for spike specific IgG ELISA assay. Dr. Chia En Lien, Dr. Meei-Yun Lin, Luke Tzu-Chi Liu, and Meng-Ju Tsai at Medigen Vaccine Biologics Corp., Taiwan for manuscript editing and revision.

